# Additional SNPs improve the performance of a polygenic hazard score for prostate cancer

**DOI:** 10.1101/2020.09.11.20188383

**Authors:** Roshan A. Karunamuni, Minh-Phuong Huynh-Le, Chun C. Fan, Wesley Thompson, Rosalind A. Eeles, Zsofia Kote-Jarai, Kenneth Muir, Artitaya Lophatananon, UKGPCS collaborators, Johanna Schleutker, Nora Pashayan, Jyotsna Batra, APCB BioResource (Australian Prostate Cancer BioResource), Henrik Grönberg, Eleanor I. Walsh, Emma L. Turner, Athene Lane, Richard M. Martin, David E. Neal, Jenny L. Donovan, Freddie C. Hamdy, Børge G. Nordestgaard, Catherine M. Tangen, Robert J. MacInnis, Alicja Wolk, Demetrius Albanes, Christopher A. Haiman, Ruth C. Travis, Janet L. Stanford, Lorelei A. Mucci, Catharine M. L. West, Sune F. Nielsen, Adam S. Kibel, Fredrik Wiklund, Olivier Cussenot, Sonja I. Berndt, Stella Koutros, Karina Dalsgaard Sørensen, Cezary Cybulski, Eli Marie Grindedal, Jong Y. Park, Sue A. Ingles, Christiane Maier, Robert J. Hamilton, Barry S. Rosenstein, Ana Vega, The IMPACT Study Steering Committee and Collaborators, Manolis Kogevinas, Kathryn L. Penney, Manuel R. Teixeira, Hermann Brenner, Esther M. John, Radka Kaneva, Christopher J. Logothetis, Susan L. Neuhausen, Azad Razack, Lisa F. Newcomb, Canary PASS Investigators, Marija Gamulin, Nawaid Usmani, Frank Claessens, Manuela Gago-Dominguez, Paul A. Townsend, Monique J. Roobol, Wei Zheng, The Profile Study Steering Committee, Ian G. Mills, Ole A. Andreassen, Anders M. Dale, Tyler M. Seibert, The PRACTICAL Consortium

**Affiliations:** Department of Radiation Medicine and Applied Sciences, University of California San Diego, La Jolla, CA, USA; Radiation Oncology, George Washington University, Washington, DC; Center for Human Development, University of California San Diego, La Jolla, CA, USA; Department of Family Medicine and Public Health, University of California, San Diego, La Jolla, CA, USA; The Institute of Cancer Research, London, SM2 5NG, UK; Division of Population Health, Health Services Research and Primary Care, University of Manchester, Oxford Road, Manchester, M13 9PL, UK; http://www.icr.ac.uk/our-research/research-divisions/division-of-genetics-and-epidemiology/oncogenetics/research-projects/ukgpcs/ukgpcs-collaborators; Institute of Biomedicine, University of Turku, Finland; Department of Applied Health Research, University College London, London, WC1E 7HB, UK; Australian Prostate Cancer Research Centre-Qld, Institute of Health and Biomedical Innovation and School of Biomedical Sciences, Queensland University of Technology, Brisbane QLD 4059, Australia; Australian Prostate Cancer Research Centre-Qld, Queensland University of Technology, Brisbane; Prostate Cancer Research Program, Monash University, Melbourne; Dame Roma Mitchell Cancer Centre, University of Adelaide, Adelaide; Chris O’Brien Lifehouse and; Department of Medical Epidemiology and Biostatistics, Karolinska Institute, SE-171 77 Stockholm, Sweden; Bristol Medical School, Department of Population Health Sciences, University of Bristol, Bristol, United Kingdom; MRC Integrative Epidemiology Unit, University of Bristol, Bristol, United Kingdom; National Institute for Health Research (NIHR) Bristol Biomedical Research Centre, University Hospitals Bristol NHS Foundation Trust and the University of Bristol, Bristol, United Kingdom; Nuffield Department of Surgical Sciences, University of Oxford, Room 6603, Level 6, John Radcliffe Hospital, Headley Way, Headington, Oxford, OX3 9DU, UK; School of Social and Community Medicine, University of Bristol, Bristol, United Kingdom; Faculty of Medical Science, University of Oxford, John Radcliffe Hospital, Oxford, United Kingdom; Faculty of Health and Medical Sciences, University of Copenhagen, 2200 Copenhagen, Denmark; SWOG Statistical Center, Fred Hutchinson Cancer Research Center, Seattle, WA, USA; Cancer Epidemiology Division, Cancer Council Victoria, 615 St Kilda Road, Melbourne, VIC 3004, Australia; Centre for Epidemiology and Biostatistics, Melbourne School of Population and Global Health, The University of Melbourne, Grattan Street, Parkville, VIC 3010, Australia; Unit of Cardiovascular and Nutritional Epidemiology, Institute of Environmental Medicine, Karolinska Institutet, SE-171 77 Stockholm, Sweden; Division of Cancer Epidemiology and Genetics, National Cancer Institute, NIH, Bethesda, Maryland, 20892, USA; Center for Genetic Epidemiology, Department of Preventive Medicine, Keck School of Medicine, University of Southern California/Norris Comprehensive Cancer Center, Los Angeles, CA 90015, USA; Cancer Epidemiology Unit, Nuffield Department of Population Health, University of Oxford, Oxford, OX3 7LF, UK; Division of Public Health Sciences, Fred Hutchinson Cancer Research Center, Seattle, Washington, 98109-1024, USA; Department of Epidemiology, Harvard T. H. Chan School of Public Health, Boston, MA 02115, USA; Division of Cancer Sciences, University of Manchester, Manchester Academic Health Science Centre, Radiotherapy Related Research, The Christie Hospital NHS Foundation Trust, Manchester, M13 9PL UK; Department of Clinical Biochemistry, Herlev and Gentofte Hospital, Copenhagen University Hospital, Herlev, 2200 Copenhagen, Denmark; Division of Urologic Surgery, Brigham and Womens Hospital, 75 Francis Street, Boston, MA 02115, USA; Sorbonne Universite, GRC n°5, AP-HP, Tenon Hospital, 4 rue de la Chine, F-75020 Paris, France; Department of Molecular Medicine, Aarhus University Hospital, Palle Juul-Jensen Boulevard 99, 8200 Aarhus N, Denmark; International Hereditary Cancer Center, Department of Genetics and Pathology, Pomeranian Medical University, 70-115 Szczecin, Poland; Department of Medical Genetics, Oslo University Hospital, 0424 Oslo, Norway; Department of Cancer Epidemiology, Moffitt Cancer Center, 12902 Magnolia Drive, Tampa, FL 33612, USA; Department of Preventive Medicine, Keck School of Medicine, University of Southern California/Norris Comprehensive Cancer Center, Los Angeles, CA 90015, USA; Humangenetik Tuebingen, Paul-Ehrlich-Str 23, D-72076 Tuebingen, Germany; Dept. of Surgical Oncology, Princess Margaret Cancer Centre, Toronto ON M5G 2M9, Canada; Department of Radiation Oncology and Department of Genetics and Genomic Sciences, Box 1236, Icahn School of Medicine at Mount Sinai, One Gustave L. Levy Place, New York, NY 10029, USA; Fundación Pública Galega Medicina Xenómica, Santiago de Compostela, 15706, Spain; http://impact.icr.ac.uk; ISGlobal, Barcelona, Spain; Channing Division of Network Medicine, Department of Medicine, Brigham and Women’s Hospital/Harvard Medical School, Boston, MA 02184, USA; Department of Genetics, Portuguese Oncology Institute of Porto (IPO-Porto), 4200-072 Porto, Portugal; Division of Clinical Epidemiology and Aging Research, German Cancer Research Center (DKFZ), D-69120, Heidelberg, Germany; Departments of Epidemiology & Population Health and of Medicine, Division of Oncology, Stanford Cancer Institute, Stanford University School of Medicine, Stanford, CA 94304 USA; Molecular Medicine Center, Department of Medical Chemistry and Biochemistry, Medical University of Sofia, Sofia, 2 Zdrave Str., 1431 Sofia, Bulgaria; The University of Texas M. D. Anderson Cancer Center, Department of Genitourinary Medical Oncology, 1515 Holcombe Blvd., Houston, TX 77030, USA; Department of Population Sciences, Beckman Research Institute of the City of Hope, 1500 East Duarte Road, Duarte, CA 91010, 626-256-HOPE (4673); Department of Surgery, Faculty of Medicine, University of Malaya, 50603 Kuala Lumpur, Malaysia; Division of Medical Oncology, Urogenital Unit, Department of Oncology, University Hospital Centre Zagreb, University of Zagreb, School of Medicine, 10 000 Zagreb, Croatia; Department of Oncology, Cross Cancer Institute, University of Alberta, 11560 University Avenue, Edmonton, Alberta, Canada T6G 1Z2; Molecular Endocrinology Laboratory, Department of Cellular and Molecular Medicine, KU Leuven, BE-3000, Belgium; Genomic Medicine Group, Galician Foundation of Genomic Medicine, Instituto de Investigacion Sanitaria de Santiago de Compostela (IDIS), Complejo Hospitalario Universitario de Santiago, Servicio Galego de Saúde, SERGAS, 15706, Santiago de Compostela, Spai; Division of Cancer Sciences, Manchester Cancer Research Centre, Faculty of Biology, Medicine and Health, Manchester Academic Health Science Centre, NIHR Manchester Biomedical Research Centre, Health Innovation Manchester, Univeristy of Manchester, M13 9WL; Department of Urology, Erasmus University Medical Center, 3015 CE Rotterdam, The Netherlands; Division of Epidemiology, Department of Medicine, Vanderbilt University Medical Center, 2525 West End Avenue, Suite 800, Nashville, TN 37232 USA; http://www.cancerresearchuk.org/about-cancer/find-a-clinical-trial/a-study-find-out-looking-gene-changes-would-be-useful-in-screening-for-prostate-cancer-profile-pilot; Center for Cancer Research and Cell Biology, Queen’s University of Belfast, Belfast, UK; NORMENT, KG Jebsen Centre, Oslo University Hospital and University of Oslo, Oslo, Norway; Department of Radiology, University of California San Diego, La Jolla, CA, USA; Department of Bioengineering, University of California San Diego, La Jolla, CA, USA; Institute of Cancer Research, Sutton, SW7 3RPm UK

## Abstract

**Abstract:** *Background:* Polygenic hazard scores (PHS) can identify individuals with increased risk of prostate cancer. We estimated the benefit of additional SNPs on performance of a previously validated PHS (PHS46).

*Materials and Method:* 180 SNPs, shown to be previously associated with prostate cancer, were used to develop a PHS model in men with European ancestry. A machine-learning approach, LASSO-regularized Cox regression, was used to select SNPs and to estimate their coefficients in the training set (75,596 men). Performance of the resulting model was evaluated in the testing/validation set (6,411 men) with two metrics: (1) hazard ratios (HRs) and (2) positive predictive value (PPV) of prostate-specific antigen (PSA) testing. HRs were estimated between individuals with PHS in the top 5% to those in the middle 40% (HR95/50), top 20% to bottom 20% (HR80/20), and bottom 20% to middle 40% (HR20/50). PPV was calculated for the top 20% (PPV80) and top 5% (PPV95) of PHS as the fraction of individuals with elevated PSA that were diagnosed with clinically significant prostate cancer on biopsy.

*Results:* 166 SNPs had non-zero coefficients in the Cox model (PHS166). All HR metrics showed significant improvements for PHS166 compared to PHS46: HR95/50 increased from 3.72 to 5.09, HR80/20 increased from 6.12 to 9.45, and HR20/50 decreased from 0.41 to 0.34. By contrast, no significant differences were observed in PPV of PSA testing for clinically significant prostate cancer.

*Conclusion:* Incorporating 120 additional SNPs (PHS166 vs PHS46) significantly improved HRs for prostate cancer, while PPV of PSA testing remained the same.

## Introduction

Optimal prostate cancer screening strategies seek to strike a balance between identifying clinically significant and potentially lethal cases that require treatment, while minimizing overdiagnosis of indolent, lower-risk cases that do not need radical treatment^1-3^. Genetic risk models have emerged as potentially useful tools that identify individuals with greater risk for being diagnosied with prostate cancer^4,5^, and so help inform if and when to initiate screening for an individual. A subset of these models called polygenic hazard scores (PHS) seeks to directly identify associations between common genetic variants and the age of diagnosis of prostate cancer by utilizing the framework of time-to-event analyses^1,6^.

We have previously reported on a PHS model for prostate cancer, PHS46, that demonstrated excellent performance in an independent test set of men from varied genetic ancestries^6^. The model incorporates genetic data of 46 unique single nucleotide polymorphisms (SNPs), and was identified through a systematic search of European men genotyped on the iCOGS chipset (Illumina, San Diego, CA). With an ever-increasing list of loci associated with prostate cancer in the literature^7-9^, we sought to determine what effect, if any, the incorporation of additional SNPs would have on the performance of PHS46.

To this end, we employed a machine-learning approach, LASSO-regularized Cox regression,^10,11^ to select SNPs from a list that included the 46 used in PHS46, as well as over 100 SNPs identified in previous analyses as having genome-wide significance for association with prostate cancer^7^. LASSO-regularized regression is an established variable selection technique in datasets with a large number of predictors and has been previously implemented as a SNP selection tool for a breast cancer polygenic risk score^12^. Performance metrics describing statistical model goodness-of-fit and clinically actionable screening utility of the LASSO-regularized PHS model for prostate cancer were compared with those achieved with PHS46 to determine the potential benefit of incorporating additional SNPs in polygenic hazard models.

## Material and Methods

### Study dataset

We obtained genotype and phenotype data from the PRACTICAL^13^ consortium for this analysis. Genotyping was performed previously on either OncoArray^13^ or iCOGS^9^ chips, and these data were previously imputed using the 1000 Genomes reference panel^14^. Missing SNP calls were replaced with the mean of the genotyped data for that SNP in the training set^1,15^. In total, data from 82,007 men with European genetic ancestry (Supplementary Table 1)^13,16^ were available for this analysis. A testing set consisting of 6,411 men (4,828 controls and 1,583 cases) enrolled in the ProtecT clinical trial was set aside for estimating the performance of the final PHS models. The data from ProtecT were chosen as the testing set because they are well characterized and were previously used for validation of PHS46^1^, allowing us to directly benchmark the performance of the updated model against previous iterations. The ProtecT trial also included biopsies of participants with elevated prostate-specific antigen (PSA) level, which permits analysis of the positive predictive value of the current clinical standard for screening, PSA testing. The remaining 75,596 individuals (25,127 controls and 50,469 cases) were used for training of the model. This first analysis was limited to men of European descent because of much greater data availability in that population, but our previous work has shown that development in Europeans can inform careful future work to assess and improve performance in other ancestries^17^.

### Model development using LASSO regularization

A list of published SNPs previously identified^1,7^ to be associated with prostate cancer was compiled. In total, 180 unique SNPs were considered for estimation within the PHS model framework. An initial screening was conducted to identify pairs of SNPs that were highly correlated (R^2^ > 0.95). For each pair of highly correlated SNPs, a univariable Cox proportional hazards model using age of diagnosis of prostate cancer as the time to event was calculated for each SNP in the pair, and the one with the larger p-value was discarded. The remaining SNPs were included as candidates for the new PHS model. The R package ’glmnet’ was used to estimate a LASSO-regularized Cox-proportional hazards model^10,11^ using age of diagnosis of prostate cancer as the time to event. The genetic data of candidate SNPs and first four European ancestry principal components were included as predictors. Controls were censored at age of last follow-up. The hyper-parameter of the LASSO-regularized model, lambda, was selected using 10-fold cross-validation^10,11^. The final form of the LASSO model was estimated at the value of lambda that minimized the mean cross-validated error.

### Characterization of LASSO-regularized PHS model

The PHS score for each of the individuals in the training and testing set was estimated as the weighted sum of the genetic counts of each of the SNPs in the PHS model, using the LASSO model coefficients as weights. Distributions of the new PHS score were compared qualitatively between training and testing groups to confirm that the model was appropriately calibrated for use in the testing set.

### Performance comparison between PHS46 and LASSO-regularized PHS

Performance in the testing set was assessed using hazard ratios (HRs) and positive predictive value (PPV), as described below. In each case, performance metrics were generated for the newly developed LASSO PHS model and for PHS46. Model coefficients for PHS46 were obtained from the literature^17^. For each performance metric, one thousand bootstrap samples of the testing set were used to generate empirical 95% confidence intervals for LASSO PHS and for PHS46. In addition, bootstrapped 95% confidence intervals were generated for the percentage change of each performance metric between the two models, using PHS46 as the 25 reference. Percent changes were deemed statistically significant if the bootstrapped 95% confidence interval did not include 0.

#### HR performance

Calibration Cox proportional hazards models were fit to the bootstrapped testing data using the PHS score as the sole predictor and the age-of-diagnosis of prostate cancer as the dependent variable. The model coefficient of this Cox regression model is referred to as the calibration factor. Next, the hazard ratio between two PHS groups, such as those in the top 5% to the middle 40% (HR95/50), is estimated as the exponential of the product of the calibration factor and the difference in mean PHS scores of each group. Hazard ratios between the top 20% to the bottom 20% (HR80/20) and the bottom 20% to the middle 40% (HR20/50) were similarly calculated. The PHS cutoffs used to define these groups were determined from the distribution of PHS in the training set controls under 70 years of age^1,15^.

A similar strategy was used to estimate the HR performance for clinically significant prostate cancer. The criteria for clinical significance were any of: Gleason score >=7, stage T3-T4, PSA concentration >= 10ng/mL, pelvic lymph nodal metastasis, or distant metastasis^18^. In this analysis, controls and low-risk (i.e., not clinically significant) cancers were censored at age of last follow-up and age of diagnosis, respectively. HRs are reported after sample-weight correction^1,17,19^ using the total number of cases and controls in the ProtecT trial to generate weighting factors.

#### PPV performance

One indicator of clinical utility of a risk-stratification approach like PHS is whether it can be used to improve the PPV of the standard clinical screening test, prostate-specific antigen (PSA). As a population-based screening study, ProtecT provides biopsy results of both cases and controls with a positive PSA result (i.e., ≥/mL). PPV performance of each model was estimated by randomly sampling individuals within the testing set with positive PSA results, while maintaining the case to control ratio of the ProtecT study (1:2). PPV is calculated as the fraction of positive PSA individuals in the top 20% (PPV80) or top 5% (PPV95) of PHS scores that had clinically significant prostate cancer.

### Cumulative incidence curves for LASSO-PHS in United Kingdom

To illustrate the utility of the LASSO PHS model in informing prostate cancer screening, cumulative incidence curves for various PHS risk groups were estimated, as described previously^20^. The age-specific general cumulative incidence curve for prostate cancer was estimated for the United Kingdom population, aged 40 to 70, using data from Cancer Research UK 2015-2017^21^. The proportion of clinically significant and non-clinically significant prostate cancer at each age was estimated using data from the Cluster Randomized Trial of PSA Testing for Prostate Cancer (CAP) trial^22^. Disease-specific cumulative incidence curves for clinically significant and non-clinically-significant prostate cancer were estimated by multiplying the general cumulative incidence curve by their respective proportions. The risk-adjusted incidence curves for individuals in the upper 5^th^ percentile and upper 20^th^ percentile were estimated by multiplying the disease-specific cumulative incidence curves by the mean value of HR95/50 and HR80/50 in the testing set, respectively. Hazard ratios were obtained using the age of diagnosis of clinically significant prostate cancer as the time-to-event and after sample-weight correction.

## Results

### SNP screening and PHS model training

Of the 180 SNPs originally considered for this study, 6 SNPs were discarded in the initial screening process of removing highly correlated SNPs. Of the 174 remaining candidate SNPs (Supplementary Table 2), 166 had non-zero LASSO model coefficients and were selected for the final PHS model (PHS166).

### PHS166 model characterization

Distributions of PHS166 score were visually consistent between training and testing sets (Supplementary Figure 1). The 20^th^, 30^th^, 70^th^, 80^th^, and 98^th^ percentiles of the reference PHS risk scores (controls in training set) were estimated as -0.411, -0.307, 0.048, 0.154, and 0.557, respectively.

### Performance comparison –PHS46 vs. PHS166

All PHS166 HR-based performance metrics showed statistically significant improvements compared to PHS46 (Table 1), for both any and clinically significant prostate cancer. The mean HR95/50 and HR80/20 values for PHS166 were roughly 36 to 55% greater than those for PHS46. For example, HR80/20 for clinically significant prostate cancer increased from 6.12 to 9.45. Similarly, HR20/50 for PHS166 was, on average, 18% lower than that for PHS46. No significant differences between models were observed in either of the PPV-based performance metrics (Table 2). Among individuals in the top 20% of risk scores with a positive PSA test, the estimated mean PPV for clinically significant prostate cancer was roughly 0.19 irrespective of the model used – indicating approximately 19% of positive PSA tests in this risk group yielded a diagnosis of clinically significant prostate cancer. By comparison, approximately 13% of all positive PSA tests resulted in a diagnosis of clinically significant prostate cancer.

**Table 1.** HR performance in testing set. Sample-weight-corrected hazard ratios are estimated for PHS166 and PHS46 in the testing set, using age-of-onset of any or clinically significant prostate cancer. The percent change for each metric is calculated using the value of PHS46 as the reference. Mean values and 95% confidence intervals are reported.

| Type of cancer | HR | PHS46 | PHS166 | Change (%) |
| --- | --- | --- | --- | --- |
| Any | HR95/50 | 3.29 [2.73,3.77] | 4.45 [3.68,5.06] | 36 [18,53] |
|  | HR80/20 | 5.15 [3.92,6.18] | 7.85 [6.04,9.33] | 53 [25,78] |
|  | HR20/50 | 0.44 [0.40,0.49] | 0.37 [0.33,0.40] | -18 [-25,-10] |
| Clinically Significant | HR95/50 | 3.72 [2.89,4.43] | 5.09 [3.84,6.05] | 37 [13,59] |
|  | HR80/20 | 6.12 [4.18,7.67] | 9.45 [6.17,11.79] | 55 [17,88] |
|  | HR20/50 | 0.41 [0.35,0.47] | 0.34 [0.29,0.39] | -18 [-28,-9] |

**Table 2.** PPV performance in testing set. Positive predictive value (PPV) of PSA testing for clinically significant prostate cancer using top 5% (PPV95) and top 20% (PPV80) cutoffs of PHS166 and PHS46 risk scores. The percent change for each metric is calculated using the value of PHS46 as the reference.

| PPV | PHS46 | PHS166 | Change (%) |
| --- | --- | --- | --- |
| **PPV95** | 0.227 [0.159,0.292] | 0.239 [0.171,0.305] | 6.3 [-25.5,32.1] |
| **PPV80** | 0.192 [0.155,0.231] | 0.187 [0.150,0.222] | -2.8 [-16.3,9.9] |

### Cumulative incidence curves for PHS166 in United Kingdom

Cumulative incidence curves for clinically significant and non-clinically significant prostate cancer for the upper 5^th^ percentile (>95^th^ percentile) and upper 20^th^ percentile (>80^th^ percentile) of PHS166 scores in the United Kingdom demonstrated expected stratification of prostate cancer risk (Figure 1).

**Figure 1.**
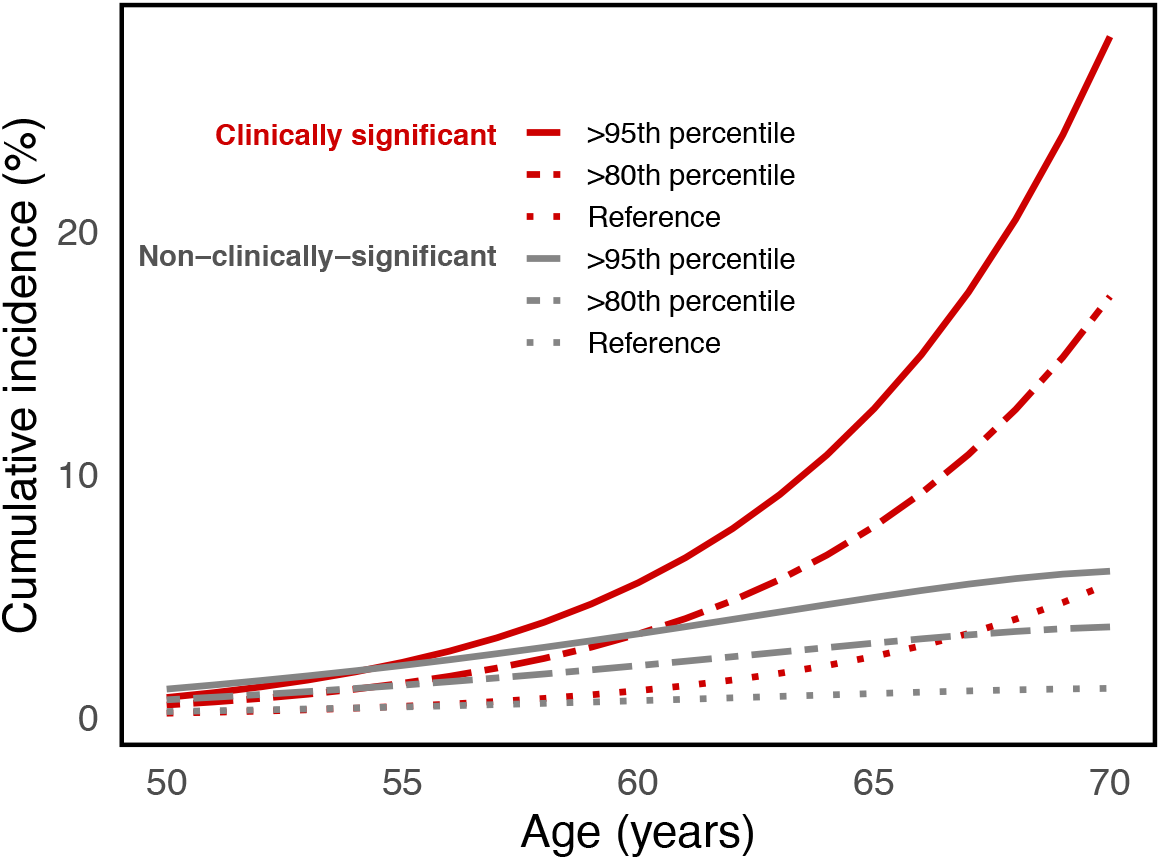
Cumulative incidence curves for PHS166. Risk-adjusted cumulative incidence curves for the upper 5^th^ percentile (>95^th^ percentile) and upper 20^th^ percentile (>80^th^ percentile) of PHS166 scores for clinically significant and non-clinically-significant prostate cancer. Reference curves representing the population average cumulative incidence (i.e., unadjusted for genetic risk).

## Discussion

Using a machine-learning, LASSO-regularized Cox framework, we identified 166 SNPs to be included in a polygenic hazard model (PHS166) for association with age of diagnosis of prostate cancer in men of European genetic ancestry. When compared to the original PHS, consisting of 46 SNPs, PHS166 demonstrated substantially improved HR performance. For example, the HR for clinically significant prostate cancer comparing the upper and lower quintiles of genetic risk increased by 56% when using PHS166. No significant improvements were found in the PPV of PSA testing when using PHS to stratify risk.

Increased separation in hazard rates between PHS risk groups may allow for more nuance in clinical decision making in certain scenarios. Accurate identification of low, intermediate, and 10 high PHS risk groups in prostate cancer may help in decisions of when (or if) to initiate screening as well as possibly improving the interpretation of the disease screens^23^. Targeting screening to men in the upper percentiles of polygenic risk as opposed to those in the lowest risk group may reduce the proportion of overdiagnosed indolent cancers from 43% to 19%^24,25^. Risk stratification achieved here by PHS166 is similar or better than commonly used clinical tools for diseases such as breast cancer, diabetes, and cardiovascular disease^23,26-28^. Clinically meaningful risk stratification is illustrated by the estimated cumulative incidence curves in Figure 1. This effect is particularly pronounced for clinically significant disease because of the increased proportion of clinically significant cases observed at older ages^2,20,22^.

The lack of improvement in PPV in this study may suggest a “performance plateau” when using PHS to define broad risk categories for certain clinical applications. A similar effect has been previously described for prostate cancer polygenic models, in the context of using risk scores to discriminate prostate biopsy outcomes^29^. Some of the precision in a score may also be diluted in broad clinical applications. The PPV analysis here is applied to participants in the ProtecT trial, which enrolled men aged 50 to 69 years, and screening in the trial was offered irrespective of underlying genetic risk^2^. Further investigation is needed to learn whether timing screening according to genetic risk might better leverage the superior HR performance of PHS166 risk score to improve the PPV of PSA testing.

LASSO frameworks have been used to identify SNPs for polygenic risk scores of several phenotypes, including fracture risk^30^, type 2 diabetes^31^, and breast cancer^12^. In this work, we have extended the application of LASSO to select SNPs in a polygenic hazard model of prostate cancer from a list of candidates previously identified through logistic and time-to-event analysis. Simulation studies^11^ have suggested that LASSO provides more robust estimates than stepwise selection in cases with both a few large effects, as well as many small effects. As new prostate cancer associated variants are discovered, this framework can be easily implemented to develop updated polygenic hazard models.

One limitation of PHS166 is that it was entirely developed and tested in European men. However, a well-vetted, well-tested PHS model for men of European genetic ancestry can be used as a starting block for developing models for other genetic ancestries, where large-scale databases are often more scarce, as has been shown for PHS46^17,32^. Furthermore, some of the SNPs selected for incorporation into PHS166 were originally discovered in analyses that included men from the ProtecT testing set. Therefore, the improvements in HRs observed for PHS166 may be somewhat overestimated. However, this bias is likely small, given that the testing set was only a small fraction of the data used in prior discovery analyses, and the ProtecT data were not used to calculate SNP weights in PHS166. In addition, this study uses age of diagnosis as the time-to-event variable, and any preceding period of undiagnosed disease is unknown. Hypothetical perfect measurement of age of onset would likely further improve performance of the PHS model.

In conclusion, we applied a machine-learning, LASSO-regularized Cox regression framework to develop a larger PHS that includes 166 previously discovered SNPs. When comparing the performance of PHS166 to the original model, PHS46, we found that incorporating 120 more SNPs significantly improved HRs for clinically significant prostate cancer. However, incorporating more SNPs did not improve on the ability of PHS46 to inform the PPV of PSA testing in the ProtecT dataset, perhaps illustrating a plateau effect and/or dilution of risk stratification in a broad clinical application.

## Data Availability

The data used in this work were obtained from the Prostate Cancer Association Group to Investigate Cancer Associated Alterations in the Genome (PRACTICAL) consortium.
Readers who are interested in accessing the data must first submit a proposal to the Data Access Committee. If the reader is not a member of the consortium, their concept form must be sponsored by a principal investigator (PI) of one of the PRACTICAL consortium member studies. If approved by the Data Access Committee, PIs within the consortium, each of whom retains ownership of their data submitted to the consortium, can then choose to participate in the specific proposal. In addition, portions of the data are available for request from dbGaP (database of Genotypes and Phenotypes) which is maintained by the National Center for Biotechnology Information (NCBI): https://www.ncbi.nlm.nih.gov/gap/?term=Icogs+prostatehttps://www.ncbi.nlm.nih.gov/gap/?term=Icogs+prostate.
Anyone can apply to join the consortium. The eligibility requirements are listed here: http://practical.icr.ac.uk/blog/?page_id=9. Joining the consortium would not guarantee access, as a proposal for access would still be submitted to the Data Access Committee, but there would be no need for a separate member sponsor. Readers may find information about application by using the contact information below:
Rosalind Eeles
Principal Investigator for PRACTICAL
Professor of Oncogenetics
Institute of Cancer Research (ICR)
Sutton, UK
URL: http://practical.icr.ac.uk
Tel: ++44 (0)20 8722 4094

## Ethics Statement

All contributing studies were approved by the relevant ethics committees and performed in accordance with the Declaration of Helsinki; written informed consent was obtained from the study participants. The present analyses used de-identified data from the PRACTICAL consortium and have been approved by the review board at the corresponding authors’ institution.

## Data Availability Statement

The data used in this work were obtained from the Prostate Cancer Association Group to Investigate Cancer Associated Alterations in the Genome (PRACTICAL) consortium. Readers who are interested in accessing the data must first submit a proposal to the Data Access Committee. If the reader is not a member of the consortium, their concept form must be sponsored by a principal investigator (PI) of one of the PRACTICAL consortium member studies. If approved by the Data Access Committee, PIs within the consortium, each of whom retains ownership of their data submitted to the consortium, can then choose to participate in the specific proposal. In addition, portions of the data are available for request from dbGaP 10 (database of Genotypes and Phenotypes) which is maintained by the National Center for Biotechnology Information (NCBI): https://www.ncbi.nlm.nih.gov/gap/?term=Icogs+prostatehttps://www.ncbi.nlm.nih.gov/gap/?term=Icogs+prostate.

Anyone can apply to join the consortium. The eligibility requirements are listed here: http://practical.icr.ac.uk/blog/?page_id=9. Joining the consortium would not guarantee access, as a proposal for access would still be submitted to the Data Access Committee, but there would be no need for a separate member sponsor. Readers may find information about application by using the contact information below:

Rosalind Eeles

Principal Investigator for PRACTICAL Professor of Oncogenetics Institute of Cancer Research (ICR) Sutton, UK URL: http://practical.icr.ac.uk Tel: ++44 (0)20 8722 4094

